# Validation of Dried Blood Spots for Capturing Hepatitis C Virus Diversity for Genomic Surveillance

**DOI:** 10.1101/2023.07.06.23292160

**Authors:** Damien C. Tully, Karen A. Power, Jacklyn Sarette, Thomas J. Stopka, Peter D. Friedmann, P. Todd Korthuis, Hannah Cooper, April M. Young, David W. Seal, Ryan P. Westergaard, Todd M. Allen

## Abstract

Dried blood spots (DBS) have emerged as a promising alternative to traditional venous blood for HCV testing. However, their capacity to accurately reflect the genetic diversity of HCV remains poorly understood. We employed deep sequencing and advanced phylogenetic analyses on paired plasma and DBS samples to evaluate the suitability of DBS for genomic surveillance. Results demonstrated that DBS captured equivalent viral diversity compared to plasma with no phylogenetic discordance observed. The ability of DBS to accurately reflect the profile of viral genetic diversity suggests it may be a promising avenue for future surveillance efforts to curb HCV outbreaks.

## BACKGROUND

Hepatitis C virus (HCV) is a significant public health problem worldwide, affecting over 50 million people, and is a leading cause of chronic liver disease, cirrhosis, and hepatocellular carcinoma. Accurate and reliable diagnosis of HCV infection is crucial for treatment and prevention. The World Health Organization’s (WHO) goal of eliminating HCV as a public health threat by 2030 will require the availability of affordable, simple point-of-care tests. The current testing paradigm requires both detection of HCV antibodies to first diagnose any previous exposure, with subsequent HCV RNA testing to confirm active infection. This may often require multiple visits where reflex testing is not available, and thus a single visit is now advocated to reduce loss to follow-up. Another barrier to testing is the stigma and lack of trust exhibited by people who inject drugs (PWID) due to prior experiences in the healthcare system. A practical difficulty is that PWID commonly experience venous degradation as a complication of prolonged injection, making it a major barrier to obtaining blood samples by venipuncture. Moreover, there was a precipitous decline in HCV antibody testing and treatment initiation during the early phases of the COVID-19 pandemic [1]. Some of these gaps in care could have been alleviated using a viable alternative to serum collection such as collection via a fingerpick. Dried blood spots (DBS) sampling involves collecting a small volume of blood on filter paper and storing it at room temperature until analysis. DBS samples are easy to collect, transport, and store, and they can be used for a wide range of diagnostic tests, including ELISA and PCR-based virus detection for SARS-CoV-2, HIV, hepatitis B virus (HBV) and HCV. DBS has several advantages over traditional diagnostic approaches, such as reduced sample volume, simplified sample collection and transport, and lower costs. DBS collections are stable at room temperature for a prolonged period making it an appealing option in low resource settings. This method of sample collection also allows for reflex virologic testing without the need of the person returning for resampling. Several studies have investigated the use of DBS for HCV testing and have reported comparable diagnostic performance to traditional plasma or serum samples [2]. However, no previous study has demonstrated the feasibility of using DBS to examine HCV genetic diversity critical to molecular surveillance and to recapitulate known transmission networks. Here, we examined the feasibility of using DBS to capture HCV sequence diversity, and when paired with deep sequencing to accurately infer the phylogenetic profile of the underlying viral population.

## MATERIALS & METHODS

### Ethics statement

All study procedures were approved by the Institutional Review Board of Massachusetts General Hospital.

### Study Participants

Specimens were taken from the Rural Opioid Initiative (ROI) study which is a cross-sectional survey of PWID in rural areas affected by the opioid crisis in the United States. For a more detailed description on the consortium see Jenkins et al [3]. A range of specimens were selected to accommodate multiple genotypes and a range of viral loads (13,800 – 13,400,000 IU/ml).

### Viral RNA extraction and PCR Amplification

#### DBS

DBS were prepared by pipetting 50ul of plasma onto each of 5 circles of a Whatman 903 card. Cards were allowed to air-dry in a flat position at ambient temperature for 4-6 hours and were packaged to simulate transport at room temperature. Two DBS per subject were cut into thin strips, placed into a 2ml o-ring tube and incubated with Buffer RLT plus β-mercaptoethanol at 37°C for 1 hour on an orbital shaker at 1000 rpm. RNA isolation proceeded per RNeasy Mini Kit protocol (Qiagen).

#### Plasma

RNA was isolated from 140 μl of plasma using the QIAamp Viral RNA Mini Kit (Qiagen, Hilden, Germany). A one step RT-PCR reaction was performed on all samples to amplify a segment at the E1/E2 junction of the HCV genome which contains the hypervariable region 1 (HVR1). The first round RT-PCR primers consisted of an Illumina adapter specific portion, a sample specific barcode segment, and an HCV HVR specific primer segment, F1-GTGACTGGAGTTCAGACGTGTGCTCTTCCGATCT-NNNNNNNNNN-GGA-TAT-GAT-GAT-GAA-CTG-GT and R1-ACA-CTC-TTT-CCC-TAC-ACG-ACG-CTC-TTC-CGA-TCT-NNNNNNNNNN-ATG-TGC-CAG-CTG-CCG-TTG-GTG-T at a final concentration of 4 pM amplified using Superscript III RT/Platinum Taq DNA Polymerase High Fidelity with the following conditions: cDNA synthesis for 30 minutes at 55°C, followed by heat denaturation at 95°C for 2 minutes, the PCR amplification conditions were 40 cycles of denaturation (94°C for 10 seconds), annealing (55°C for 10 seconds) and extension (68°C for 10 seconds) with a final extension at 68°C for 5 minutes. Amplified products were run on a 1% agarose gel and either PCR purified with the Qiaquck PCR purification kit (Qiagen) or gel extracted and purified using the PureLink quick gel extraction kit (Invitrogen). A second round limited cycle PCR (94°C for 2 minutes, (94°C for 15 sec; 55°C for 30 sec; 68°C for 30 sec) x 8 cycles, 68°C for 5 minutes) is performed to add barcode specific indexes and sequencing specific adapters and primers to each sample to allow for multiplexing as well as internal controls for cross-contamination. Negative controls were introduced at each stage of the procedure and all PCR procedures were performed under PCR clean room conditions using established protocols. Indexed samples are 0.7X SPRI bead (Beckman Coulter, Inc) purified two times to remove excess primer dimer and short fragments that can interfere with the sequencing process.

#### Deep Sequencing and Analysis

Reads were automatically de-multiplexed and subject to stringent cleaning and quality control procedures as previously outlined [4]. A *de novo* consensus assembly was generated using the Vicuna v1.1 algorithm with automated computational finishing and annotation of *de novo* viral assemblies performed using V-FAT v1.1. LoFreq v2.1.5 [5] was used as a high quality and highly sensitive tool to call single nucleotide variants (SNVs). The genomic nucleotide diversity (π) based on all reads was calculated as the average number of nucleotide differences per base pair.

#### HCV Transmission Network Analyses

Paired-end reads for each sample were upload to the CDC’s GHOST system [6] for quality control and to analyze for the presence of transmission links using a previously empirically defined hamming distance based threshold of 0.037 [7].

#### Phylogenetic and compartmentalization analyses

All *de novo* consensus sequences were aligned using MAFFT v7.470 and a maximum likelihood phylogenetic tree was reconstructed using IQ-TREE v 2.1 with the best-fit model of nucleotide substitution according to the Bayesian Information Criterion (BIC). Phyloscanner v1.8 [8] was used to reconstruct deep sequencing phylogenies from reads and identify probable transmission clusters. Viral populations were assessed for compartmentalization using two tree-based methods: the Slatkin-Maddison test and a modified Slatkin-Maddison test which reduces spurious compartmentalization detection in trees with large numbers of sequences. This test was performed on phylogenetic trees in which identical reads are collapsed to a single read giving a focus on diverse populations. Phylogenetic trees were considered compartmentalized if 10,000 permutations of the standard Slatkin-Maddison test and 50,000 permutations of the structured-Slatkin-Maddison test yielded a *P* value of <0.05. All tests were performed in HyPhy v2.5.32 [9].

## RESULTS

To determine the capacity of DBS to replace plasma to conduct HCV genetic linkages for outbreak investigation analyses we compared the HCV sequences generated from each sample source to a panel of 14 HCV infected subjects. We PCR amplified and deep sequenced paired plasma and DBS samples obtained from 14 subjects with HCV infection who were either infected with genotype 1a or 3a. The subjects had plasma viral loads ranging from 13,800 to 13,400,000 IU/ml allowing us to test a broad range of input HCV viral loads (**Table 1**). Deep sequencing of paired DBS and plasma samples revealed no significant differences in terms of sequencing coverage (**Table 1**; *P* = 0.22) and both sample types were sequenced to equitable depth.

**Table 1.** Overview of study subject characteristics and sequencing depth as measured by the number of reads. ND refers to not determined

| Sample Name | Genotype | Viral Load (IU/ml) | Median Coverage |  | Coverage (Standard Deviation) |  |
| --- | --- | --- | --- | --- | --- | --- |
|  |  |  | Plasma | DBS | Plasma | DBS |
| Sample6001 | 1a | 10,900,000 | 20064 | 15836 | 964 | 383 |
| Sample6029 | 1a | 3,470,000 | 19511 | 26550 | 679 | 415 |
| Sample6050 | 1a | 58,800 | 18451 | 27568 | 832 | 747 |
| Sample3013 | 1a | 47,500 | 12660 | 23194 | 589 | 667 |
| Sample3048 | 1a | 13,800 | 11158 | 14630 | 771 | 3913 |
| Sample2026 | 1a | 131,000 | 16876 | 15785 | 1388 | 297 |
| Sample1045 | 1a | 3,820,000 | 20457 | 30171 | 1168 | 920 |
| Sample1152 | 1a | 97,700 | 21977 | 21312 | 971 | 11968 |
| Sample1168 | 3a | 13,400,000 | 15581 | 30381 | 789 | 1093 |
| Sample2085 | 1a | ND | 21774 | 27257 | 4188 | 7485 |
| Sample3095 | 1a | ND | 59772 | 56862 | 14066 | 5264 |
| Sample2308 | 1a | ND | 28936 | 36963 | 2170 | 5706 |
| Sample5079 | 1a | ND | 45861 | 55075 | 3334 | 3828 |
| Sample1059 | 3a | ND | 56289 | 39221 | 4169 | 2516 |

Maximum-likelihood phylogenetic analysis of the consensus sequences showed that DBS samples were phylogenetically indistinguishable from those derived from plasma with 100% identify observed in all but 3 samples which shared 99.67% identity **(Figure 1a)**. To test for any bias associated with DBS versus plasma source we tested for differences in viral diversity and compartmentalization using the deep sequencing data. Intra-host sequence diversity was not significantly different in any individual when tested **(Figure 1b)** while the variant frequencies found in DBS were highly concordant from matching plasma (R^2^ = 0.97; **Figure 1c)**.

**Figure 1.**
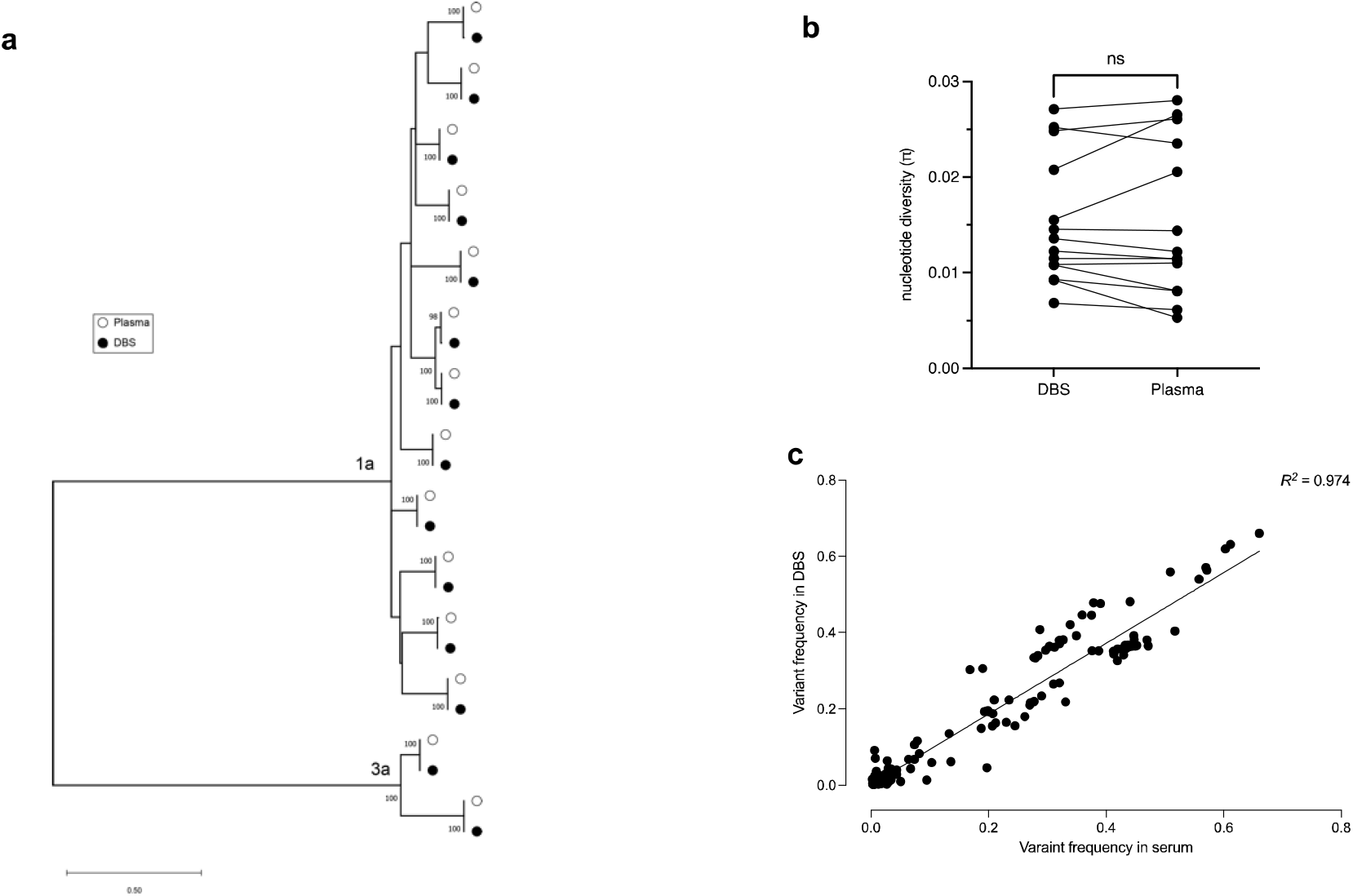
(a) Maximum-likelihood tree of DBS (closed black circle) and plasma (open white circle) derived consensus sequences. (b) overall intra-host nucleotide diversity within each sample. Lines denote paired samples between DBS and Plasma (c) Correlation of variant frequencies observed for SNVs candidates detected with deep sequencing of DBS and plasma.

To investigate whether viruses from DBS had any evidence of compartmentalization we implemented two phylogenetic-based approaches using the Slatkin-Maddision test and a recently modified Structured Slatkin-Maddison test which reduces spurious detection of compartmentalization when dealing with large number of sequences [10]. Trees were considered compartmentalized if both tests yielded a P-value <0.05. Among the 14 samples we found that none of the samples showed statistical evidence using both tests for compartmentalization in DBS (**Supplementary Table 1**). Finally, using the Global Hepatitis Outbreak and Surveillance Technology (GHOST) platform, a CDC viral hepatitis web-based system to identify HCV genetic linkage between individuals, we demonstrated that paired DBS and plasma samples all linked as expected as individual clusters (**Supplementary Figure 1**). Within this dataset we were able to reconstruct a known transmission network between two infected individuals (depicted as a cluster of size 4 in **Supplementary Figure 1**). Intra-host genetic diversity relatedness across all sample pairs were visualized using k-step networks where despite showing a broad range of heterogeneity the structure of intra-host HCV HVR1 populations was highly related (**Supplementary Figure 2**). Network analyses demonstrated that DBS and plasma from both these infected individuals were intermingled and below a previously validated genetic relatedness threshold [7]. To further investigate the phylogenetic relationship of DBS and plasma derived sequences we performed deep sequence phylogenetics using Phyloscanner [8]. These studies demonstrated that DBS and plasma-derived sequence reads were very intermingled throughout the tree with a negligible subgraph distance of 1.0e-6 substitutions per site separating pairs.

## DISCUSSION

Genomic surveillance efforts can be used to study the transmission and evolution of pathogens such as HCV and has increasingly been recognized as an integral component to guide public health interventions aimed at interrupting transmission [11,12]. Owing to its high mutation rate HCV can quickly adapt leaving a genomic footprint which provides valuable empirical information on its transmission history when assessed by phylogenetic means. DBS offers an alternative to established phlebotomy requiring a smaller volume of blood than plasma or serum and it mitigates the need for cold-chain refrigeration efforts. In addition, DBS specimens once completely dried are considered noninfectious and nonhazardous allowing them to be shipped and transported at ambient temperatures. The provision of such alternative diagnosis approaches to low income, remote and hard-to-reach populations may help close the gaps in eliminating HCV by 2030. Therefore, using DBS as an alternative means for genomic sequencing efforts could empower HCV surveillance initiatives and enable point of care genomic analysis.

Several studies evaluating the diagnostic accuracy of DBS testing for HCV testing have demonstrated that they are a reliably sensitive tool to diagnose viremic infection [13], discriminate reinfection from relapse [14] and monitor antiviral resistance [15]. However, DBS have not been completely validated by deep sequencing with respect to how well they capture HCV genomic diversity at the intra-host and inter-host levels. The present study demonstrates accurate consensus-level HCV sequence determination with DBS, with consensus level sequences detected with >99.7% accuracy when compared to plasma and phylogenetically inseparable. We also report concordant intra-host diversity and no significant evidence of compartmentalization of DBS and plasma-derived sequences. Thus, the virus populations between DBS and plasma are well equilibrated and there is no evidence to support that DBS lacks the sensitivity in its ability to detect viral lineages. Similarly, an evaluation of the intra-host nucleotide diversity revealed that single nucleotide variants (SNVs) were accurately detected at a frequency of >2% with similar frequencies.

In conclusion, investigation of a range of sample viral loads from two major genotypes reveals that robust phylogenetic analysis incorporating intra-host diversity can be successfully performed using DBS samples. These results and the practical advantages afforded by DBS supports their use in augmenting molecular surveillance activities and highlights their future importance in focusing on elimination efforts of HCV.

## Data Availability

All data produced in the present study are available upon reasonable request to the authors

## Acknowledgements

This work was supported by the National Institute of Drug Abuse (NIDA) grant U24DA044801. Data is based upon data collected and/or methods developed as part of the Rural Opioid Initiative (ROI), a multi-site study with a common protocol developed collaboratively by investigators at eight research institutions and at the National Institute of Drug Abuse (NIDA), the Appalachian Regional Commission (ARC), the Centers for Disease Control and Prevention (CDC), and the Substance Abuse and Mental Health Services Administration (SAMHSA). Primary data collection was supported by grants UG3DA044798, UG3DA044830, UG3DA044831, UG3DA044826 co-funded by NIDA, ARC, CDC, and SAMHSA. The authors thank the other ROI investigators and their teams, the ROI Executive Steering Committee chair, Dr. Holly Hagan, the NIDA Science Officer, Dr. Richard Jenkins, and, particularly, the participants of the individual ROI studies for their valuable contributions. A full list of participating ROI investigators and institutions can be found on the ROI website at http://ruralopioidinitiative.org/studies.html.

## Supplementary Information

**Supplementary Table 1.** Summary of the genetic compartmentalization results to determine the relationship between DBS-derived and plasma-derived HCV sequences. Bolded values indicate statistically significant compartmentalization as determined by each method.

| Sample Name | Structured Slatkin-Maddison value | Standard Slatkin-Maddison value |
| --- | --- | --- |
| Sample6001 | 0.941 | 0.781 |
| Sample6029 | 1 | 1.000 |
| Sample6050 | 0.688 | **0.015** |
| Sample3013 | 1 | 0.688 |
| Sample3048 | 0.979 | 0.857 |
| Sample2026 | 0.993 | 0.993 |
| Sample1045 | 0.824 | 0.246 |
| Sample1152 | 0.593 | 0.325 |
| Sample1168 | 1 | 1.000 |
| Sample2085 | 0.907 | 0.998 |
| Sample3095 | 0.850 | 0.911 |
| Sample2308 | 0.955 | 0.955 |
| Sample5079 | 0.943 | 0.975 |
| Sample1059 | 0.077 | **0** |

**Supplementary Figure 1.**
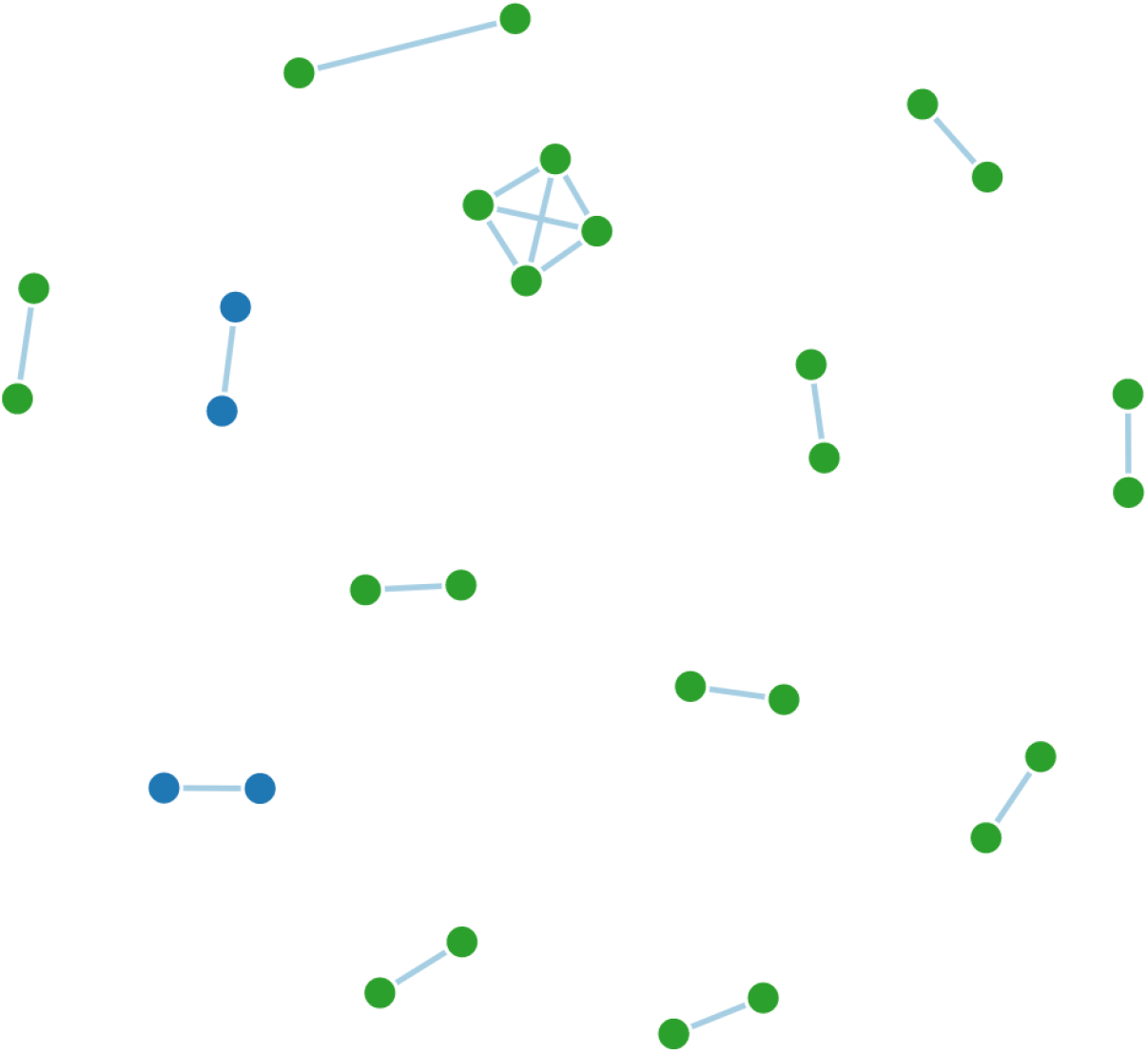
Transmission network analysis of paired DBS and plasma samples. Each node represents a unique individual’s plasma or DBS sample. A link is drawn if the minimal hamming distance between sequences from two samples is smaller than the established genetic distance threshold. The cluster of size 4 reflects a known transmission network between two infected individuals. Clusters are color coded by genotype with dark blue indicating genotype 3 and dark green indicating genotype 1a samples.

**Supplementary Figure 2.**
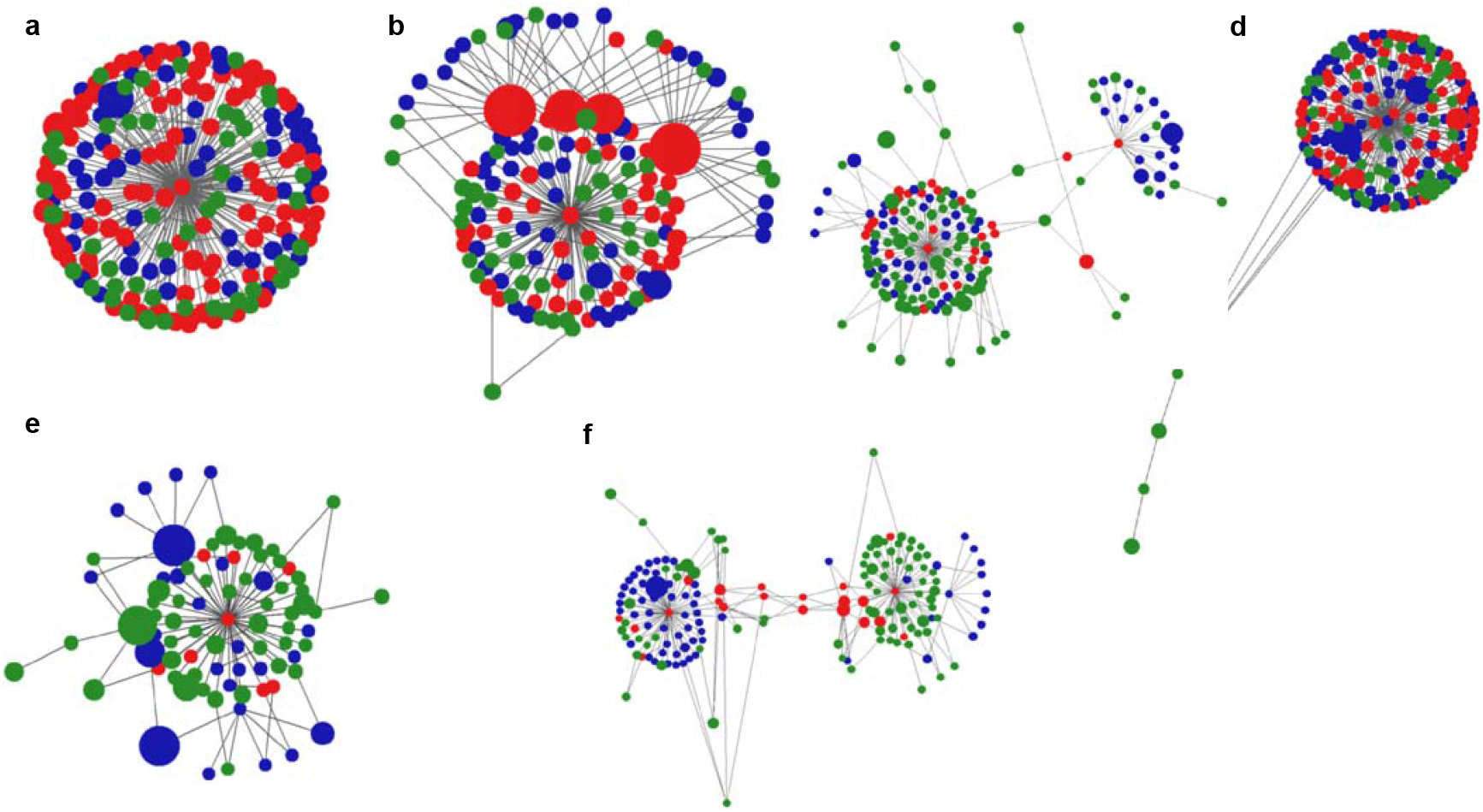
K-step network of the HCV variants found in 6 representative paired samples. The k-step network contains all the possible minimum spanning trees and is used to visualize the genetic relatedness of all haplotypes present in a sample. Each node represents a unique haplotype and the diameter of the node is proportional the haplotype frequency. Links between adjacent nodes belong to the union of all minimum spanning trees. Green/blue color identifies the source of each haplotype while red represent those haplotypes that are shared by both samples.

## Notes

### Competing Interest Statement

The authors have declared no competing interest.

